## Supplementary material for "Remdesivir as a tool to relieve hospital care systems stressed by COVID-19: A modelling study on bed resources and budget impact"

**Appendix**

**Supplementary Figure 1**: New SARS-CoV-2 infections per day relative to the number of diagnosed infections observed (A), COVID-19-related deaths (B), hospital admissions (C), hospital beds occupied (D), ICU admissions (E) and ICU beds occupied (F) estimated with the model for current care in France versus observed data between 14/03/2020 and 18/11/2020

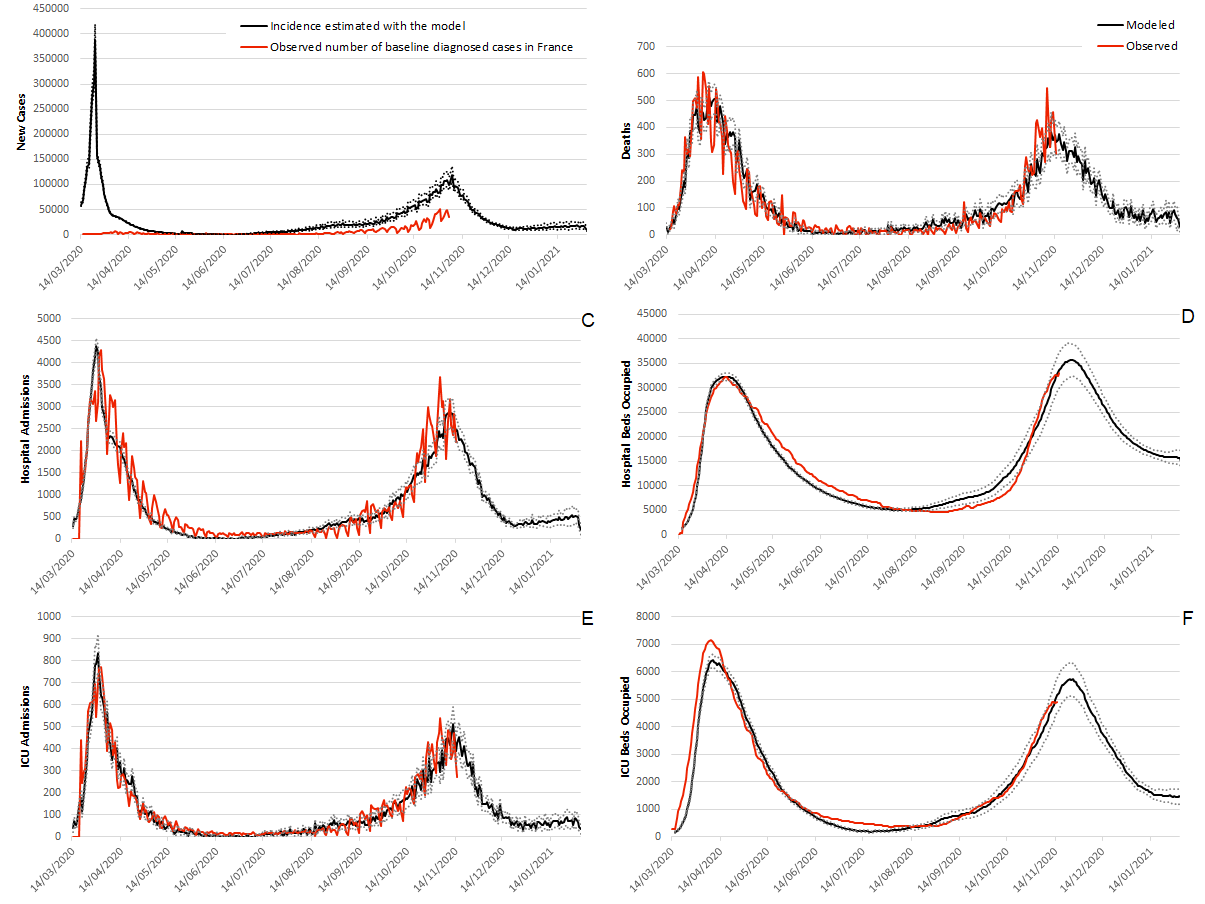

*** The discrepancy between diagnosed cases and estimated incidence is in agreement with what was observed in France and what was measured by incidence studies from Santé Publique France. Many mild cases remained undiagnosed, mostly due to the limitations of testing capacities.**

**Supplementary Figure 2**: ICU beds occupied per 100,000 individuals (A), absolute decrease in ICU bed occupancy per 100,000 individuals with the use of dexamethasone for ICU patients (B) and relative reduction in bed occupation with the use of dexamethasone for ICU patients relative to the current standard of care in France (C) between 01/08/2020 and 01/02/2021.

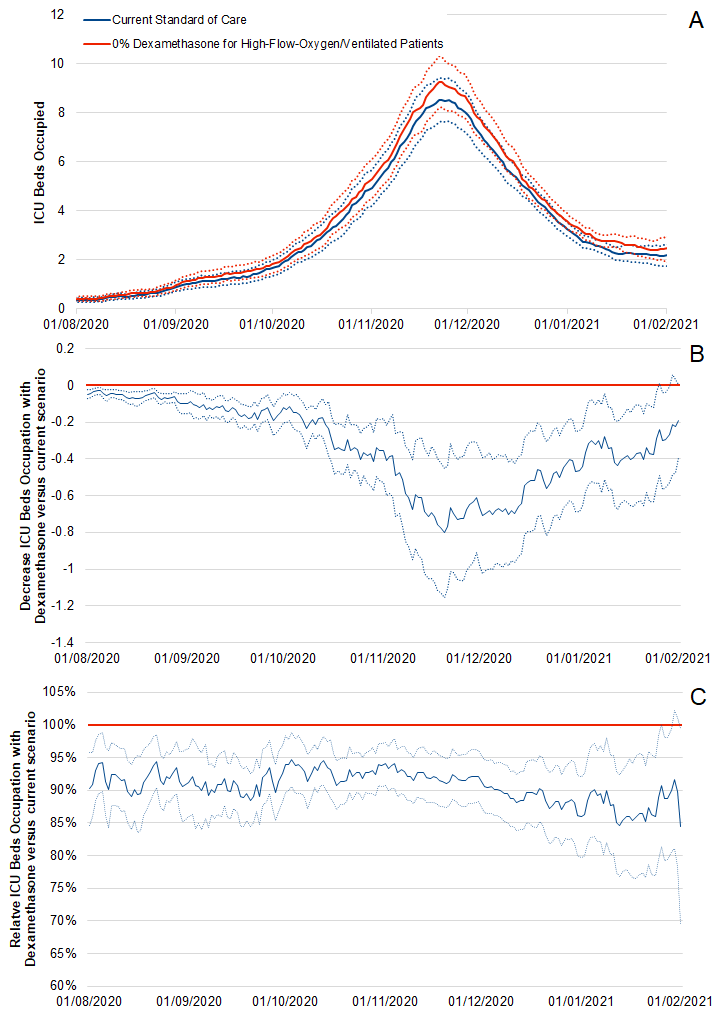

**Supplementary Table 1. Summary of main model parameters.**

| **Parameters** | **Value** | **Source** |
| --- | --- | --- |
| **Individuals’ characteristics** |  |  |
| Family structure (%) |  | Insee, 2020 (7) |
| Single | 37% |  |
| Couples with children | 27% |  |
| Couples without children | 27% |  |
| Single parents with children | 9% |  |
| Age structure (categorized by 5-year age groups) |  | Insee, 2020 (7) |
| Prevalence of conditions associated with increased risk of death from COVID-19 (i.e., obesity, diabetes, chronic cardiac diseases, and chronic respiratory diseases) | Estimates per 10-year age groups and sex | (8,17,33,34) |
| **Social contacts** |  |  |
| School class size (average) | 30 | Assumption |
| Proportion of small companies (<10 employees) | 18.40% | Insee, 2018 (35) |
| Number of colleagues in small companies (average) | 2 | Assumption |
| Number of colleagues in bigger companies (average) | 10 | Assumption |
| Employment rate (for people aged 20 to 65 years) | 92% | Insee, 2018 (35) |
| Shopping density (per 100,000 inhabitants) | 29.3 | APUR, 2018 (36) |
| Number of shopping trips (average per week) | 1.2 | Assumption |
| Number of people met per shopping trip (average) | 5 | Assumption |
| Social network distance | 22 | (37) |
| Frequency of meeting friends (average per week) | 1 | Assumption |
| Events, i.e., museum, cinema, music and sport events (average per year) | 5.4 | French Ministry of Culture, 2018 (38) |
| Close encounters per event (average) | 5 | Assumption |
| Round trips with public transport (average per week) for workers | 5 | Assumption |
| Round trips with public transport (average per week) for non-workers | 1.7 | Assumption |
| Close encounters in public transport | 3 to 5 | Assumption, with work-related trips assumed to happen at peak times with more encounters |
| International contaminations (average, per week) | 1.8 | Based on imported cases observed in France initially (Santé Publique France) (17) |
| **SARS-CoV-2 infection characteristics** |  |  |
| Contamination risk (per min/m²) | 0.003 | Estimated through model calibration, based on daily mortality data through April 15, the cumulative number of cases diagnosed through April 15, and the assumption of a cumulative incidence (diagnosed + undiagnosed) of a 1 in 100 diagnosis rate |
| Proportion of asymptomatic/paucisymptomatic individuals who will not be diagnosed | [52%-98%] |  |
| Proportion of asymptomatic people | 25% | London Imperial College, 2020 (39) |
| Hospitalization rates | [0.1%-31.4%] | Institut Pasteur (12) |
| ICU rates (if hospitalized) | [3.4%-36.4%] | Institut Pasteur (12) |
| Mortality rates (if hospitalized) | [0.0%-42.0%] | Institut Pasteur (12) |
| Impact of comorbidities on outcomes (Hospitalization, ICU, Mortality) | Hazard ratio estimates adjusted for sex and age and other comorbidities | (13) |
| Delays (days) |  |  |
| Incubation time (average, standard deviation) | 6.4 (2.3) | (15) |
| Infection onset to diagnosis (average, standard deviation) | 2.1 (2.6) | (16) |
| Infection onset to hospital admission (average, standard deviation) | 5.8 (4.2) | Institut Pasteur (12) |
| Hospital admission to recovery or transfer to rehabilitation care (average, standard deviation) (no oxygen or low-flow oxygen) | 8.0 (1.0) | DRESS (14) |
| Hospital admission to death (average, standard deviation) (no ICU admission) | 4.0 (1.0) | DRESS (14) |
| Rehabilitation care to recovery (average, standard deviation) (no oxygen or low-flow oxygen) | 33.0 (3.3) | DRESS (14) |
| Hospital admission to ICU (average, standard deviation) (initially low-flow oxygen) | 4.0 (1.0) | DRESS (14) |
| ICU admission to ICU discharge or death (average, standard deviation) (short stay) | 16.0 (1.6) | DRESS (14) |
| ICU admission to ICU discharge or death (average, standard deviation) (long stay) | 35.0 (3.5) | DRESS (14) |
| ICU discharge to recovery or rehabilitation care (average, standard deviation) | 4.0 (1.0) | DRESS (14) |
| Infection onset to recovery (average, standard deviation) (no hospitalization) | 20.5 (6.7) | London Imperial College (39) |
| Proportion of hospitalized patients with mild or moderate disease on low-flow oxygen | 78.2% | Assumption |
| Proportion of hospitalized patients with mild or moderate disease requiring rehabilitation care | 12.4% | DRESS (14) |
| Proportion of hospitalized patients with severe disease admitted directly to the ICU | 63.0% | DRESS (14) |
| Proportion of hospitalized patients admitted to the ICU with a long stay | 15.0% | DRESS (14) |
| RT-PCR sensitivity (average) | 71% | (40) |
